# Using the sanitation safety planning tool to improve the occupational health and safety of de-sludging operators in Uganda: a protocol for a controlled before and after study

**DOI:** 10.1101/2022.07.01.22277149

**Authors:** Tonny Ssekamatte, Aisha Nalugya, Samuel Fuhrimann, Richard K. Mugambe, Winnifred K. Kansiime, Arnold Tigaiza, Doreen Nakalembe, Nishai Moodley, Fred Nuwagaba, Ceaser Kimbugwe, Jane Sembuche Mselle, Bridget Nagawa Tamale, Abdullah Ali Halage, John Bosco Isunju

## Abstract

**Background:** De-sludging operators play a critical role in ensuring access to safe sanitation services. De-sludging activities, however, increase the operators’ risk of exposure to physical, chemical, ergonomic, psychological, and biological hazards, which eventually affect their health-related quality of life (HRQoL). Despite immense exposure to occupational hazards, only a few evidence-based interventions have been implemented to improve the occupational health and safety (OHS) of de-sludging operators.

**Objective:** The proposed study intends to utilise the sanitation safety planning (SSP) tool to improve the OHS of de-sludging operators in Uganda. The study aims to generate evidence of the prevalence of exposure to occupational hazards and the associated health outcomes, knowledge, attitude and uptake of OHS measures, as well as the HRQoL of de-sludging operators. The facilitators and barriers to the promotion of OHS shall also be explored.

**Methods:** A mixed-methods study will be conducted among 356 desludging operators in the Greater Kampala Metropolitan Area. A structured questionnaire pre-loaded on the KoboCollect mobile data collection application will be used to obtain information on socio-demographic characteristics, history of work-related accidents and diseases, uptake of safety measures, and health-related quality of life. A total of 10 focus group discussions (FGDs) will be conducted among desludging operators, to understand how they cope with associated occupational hazards. In addition, 25 in-depth interviews (IDIs) will be conducted among purposively selected employers to understand the barriers and facilitators of provision and implementation of safety measures among desludging operators. Quantitative data will be analyzed using STATA version 15 while qualitative data will be transcribed verbatim and the analysis aided by the NVIVO software.

**Expected outcomes:** The study will generate evidence of the occupational health and safety of desludging operators and the effectiveness of the SSP tool. These findings will be critical in informing the design and implementation of occupational health and safety programmes among desludging operators.

## Background

Globally, a vast majority of the urban population, especially in low and middle-income countries rely on on-site sanitation technologies for human excreta disposal [1, 2]. Particularly, over 80% of the urban dwellers in sub-Saharan Africa depend on on-site sanitation technologies, such as septic tanks and pit latrines [2]. The sustainability of such technologies necessitates regular and safe emptying, and disposal of faecal sludge [2, 3].

De-sludging operators are individuals responsible for safe emptying, transportation, and disposal of faecal sludge. They play a critical role in ensuring equitable access to sanitation services, especially in low- and middle-income countries where sewer systems are less common [4]. De-sludging operators contribute to the reduction of environmental pollution and the transmission of wastewater and sanitation-related diseases (such as cholera, hepatitis E, and soil-transmitted helminth), as well as upholding human dignity by lowering human contact with excreta and wastewater. Desludging is associated with both environmental and public health benefits. However, despite these benefits, de-sludging operators face enormous occupational risks, which remain largely undocumented to support effective prevention measures [5].

There is limited literature on occupational hazards faced by de-sludging operators, and coping mechanisms, although evidence suggests physical (e.g. slips, trips, falls and radiation), ergonomic (e.g., musculoskeletal disorder), chemical, biological (such as bacteria, viruses), and psychosocial hazards (e.g., stress, bullying, and harassment) among toilet operators [5, 6]. Yet, de-sludging operators in low- and middle-income countries, such as Uganda, engage in riskier activities compared to other sanitation workers, including toilet operators. De-sludging operators engage in risky activities, such as manual de-sludging and gulping, which exposes them to microbiological, physical, and psychological hazards. These hazards are exacerbated by the poor state of sanitation facilities; lack of adequate physical access for cesspool/pit-emptying due to poor planning; and lack of proper personal protective equipment (PPE) for adequate protection of workers [7, 8]. Besides, occupational hazards are known to affect their HRQoL by aggravating the risk of pain, suffering, deterioration of mental health, and premature death [5, 6].

Despite these consequences, little is known about the occupational health and safety (OHS) of de-sludging operators in Uganda, where many workers in Uganda are unaware of their rights to a safe and healthy working environment. This study will use the SSP tool to establish the occupational hazards and available safety measures among de-sludging operators in Uganda, to inform relevant policy interventions. The SSP tool is a risk-based management tool used to systematically identify and manage health risks along the sanitation chain, and guide investment based on actual risks. This tool will also promote health benefits and minimize adverse health impacts, and provide assurance to authorities and the public on the safety of sanitation-related products and services [9].

### Theoretical model

In addition to the SSP tool, the proposed study will also make use of the health belief model (HBM) to influence OHS behaviours. The HBM is built on the perceived facets of severity, susceptibility, benefits, and barriers, as well as cues to action, and self-efficacy [10-12]. The HBM can be used to guide health promotion and disease prevention programs. This study theorizes that uptake of desired prevention behaviour is influenced by a person’s subjective perception of the risk of acquiring an occupational health-related injury or disease (perceived susceptibility); feelings on the seriousness of contracting an occupational health-related injury or disease (perceived severity); perception of the effectiveness of various actions available to reduce the threat of occupational injury or disease (perceived benefits); a person’s feelings on the obstacles to performing a recommended health action (perceived barriers); stimulus needed to trigger the decision-making process to accept a recommended health action (cues to action), and the level of a person’s confidence in one’s ability to successfully perform a behaviour (self-efficacy) [13, 14].

### Research questions and study objectives

#### Research questions

1. What is the prevalence of occupational hazards among desludging operators in cities in Uganda?
2. What is the level of knowledge, attitude and uptake of occupational health and safety measures among desludging operators in cities in Uganda?
3. What are the facilitators and barriers to promotion of occupational health and safety of desludging operators in cities in Uganda?

### Main study objective

To use the sanitation safety planning (SSP) tool to improve the occupational health and safety (OHS) of de-sludging operators in Uganda

### Specific objectives

1. To establish the prevalence of occupational hazards among desludging operators in cities in Uganda
2. To establish the knowledge, attitude and uptake of occupational health and safety measures among desludging operators in cities in Uganda
3. To understand the facilitators and barriers to the promotion of occupational health and safety of desludging operators in cities in Uganda

## Materials and methods

### Study setting

This study will be conducted in Greater Kampala Metropolitan Area (GKMA) and in the cities of Fort portal, Arua, Gulu, Jinja, Mbarara, Mbale, Lira, Soroti, Masaka and Hoima (Fig 1). Fort portal city is located in Kabarole district in the western region of Uganda. Fort portal, Arua, Mbarara, Jinja and Gulu are the newest cities, elevated from municipality to city by the Parliament of Uganda and became operational from 1 July 2020 [15]. Fort portal is located in Kabarole district and has a total population of 53,786 people, of which 51.1% are males. Arua city is located in Arua district in the northern region of Uganda, with a total population of 61,962 people. Mbarara city is located in the western region of Uganda and has a total population of 195,318, of which 49.0% are males. Jinja city is within the eastern region and has a total population of 32,771 49.2% of which are males. Gulu city is located in the northern region of Uganda, with a total population of 150,306 people. Mbale. Lira city is located in the northern region of Uganda with a total population of 99,392 of which 47.1 % are males. Masaka city is located in the central region with a total population of 103,227 of which 47.4% are males. Hoima city is located in the western region of Uganda with a total population of 100,099 of which 47.6% are males. Soroti city is a city in the Eastern Region of Uganda of which 49,685 of 48.3% are males [16]. The GKMA comprises Kampala Capital City and the neighbouring districts of Wakiso and Mukono. The area contributes a third of Uganda’s overall gross domestic product (GDP) and has 46% of all formal employment. It accounts for 10% of Uganda’s population (3.5 million) during the day [17].

**Figure 1:**
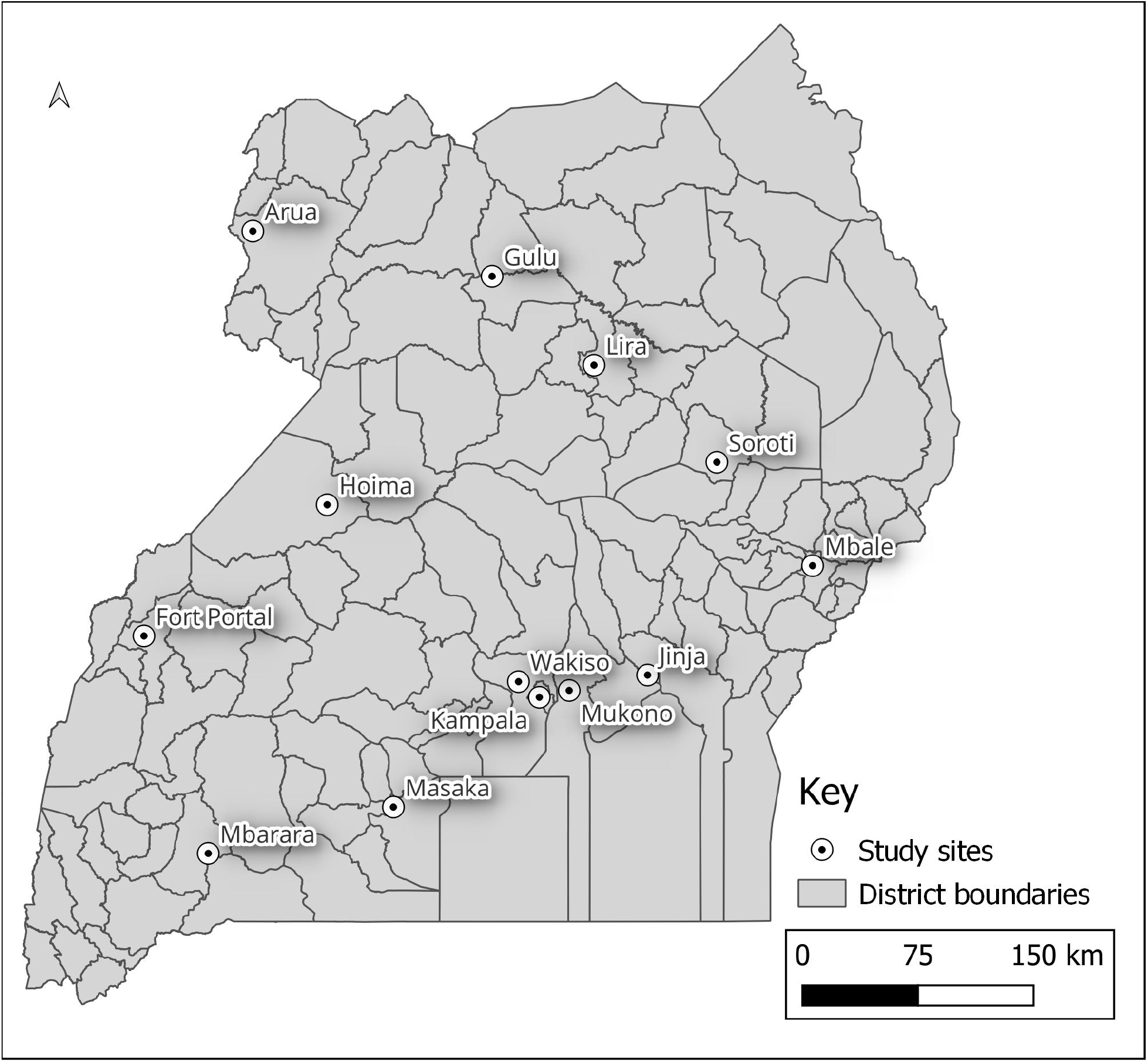
Map of Uganda showing the study sites/cities

**Figure 2:**
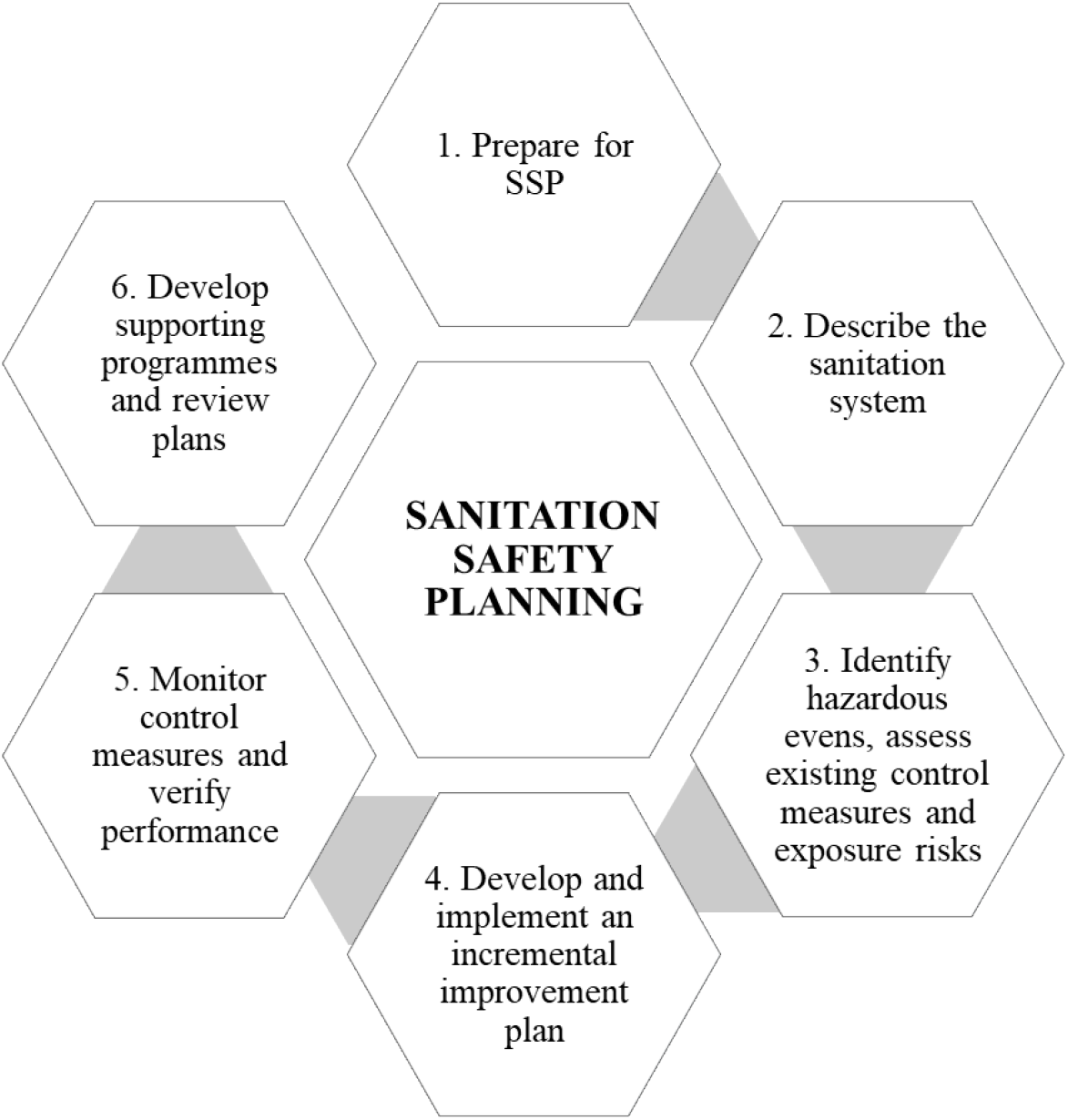
Sanitation safety planning (SSP) modules.

Although data from other regions are scarce, available statistics indicate that Kampala City has more than 83 cesspool trucks (private and institutional), 7 of which are owned by Kampala Capital City Authority, and 43 by the Private Emptiers Association Uganda and Kampala Emptiers Association. Additionally, there are 2 schools in Kampala that own and operate their trucks, while NGOs such as Community Integrated Development Initiative have recently acquired 3 cesspool emptiers (vacuum trucks) under the Kawempe Urban Poor Sanitation Improvement Project to serve Kawempe division [18]. The remaining trucks belong to a hotel, the army, the police, and two private companies [7].

### Study design, population, and eligibility criteria

This research will be a controlled before-and-after study that will utilise both qualitative and quantitative data collection methods. The study will be conducted among de-sludging operators. In order to be eligible to participate in the study, a de-sludging operator must have engaged in emptying, transportation, and disposal of feacal sludge in the above selected cities.

### Study approach

The study will be guided by the six stages of SSP. These stages include (1) preparation for SSP, (2) description of the sanitation system, (3) identification of hazardous events, assessment of existing control measures and exposure risks, (4) development and implementation of an incremental improvement plan, (5) supporting the monitoring of control measures and performance, (6) developing supporting programmes and reviewing plans. The preparatory stage will aim to establish priority areas or activities, set objectives, define the system boundary and lead organization, and assemble the SSP team. The description of the sanitation system will include mapping of the sanitation system, characterization of the waste fractions, identification of potential exposure groups, gathering compliance and contextual information, and validation of feacal sludge system description. During the identification of hazardous events, existing control measures and exposure risks will be assessed. During stage 3, a risk assessment table will be developed that includes a comprehensive list of hazards, summarized hazardous events, exposure groups and routes, existing control measures and their effectiveness, and a prioritized list of hazardous events to guide system improvements.

Stage 4 will involve the identification and consideration of options to control identified risks, use selected options to develop an incremental improvement plan, and implement the improvement plan. The project team will work with the de-sludging operators and their leadership to develop a behaviour change intervention that will aim to improve occupational health and safety through the utilisation of personal protective measures. The intervention will involve the use of the short message service (SMS) and the distribution of informative pamphlets. De-sludging operators will be contacted through one-way text messages using SMS (five SMS on occupational health and safety of up-to 160 characters will be sent to de-sludging operators bi-weekly) or social media platforms such as WhatsApp to further create awareness of occupational health and safety. All the text messages will be sent in the language preferred by the desludging operators. In addition, informative pamphlets on occupational health and safety will be customized and distributed. Each de-sludging operator will be provided with an informative pamphlet on occupational health and safety. The pamphlet will be written in different languages (with illustrations or diagrams) to accommodate both a literate and non-literate audience. An electronic version of the pamphlet will also be emailed to desludging operators, for those with email addresses or social media accounts on WhatsApp, Facebook, Instagram, and other similar platforms. Informative pamphlets will be distributed after the baseline data collection. During stage 5, the feacal sludge management associations will be supported, alongside the local authority to define and implement operational monitoring, verify system performance, and audit the system.

The controlled before-and-after study design to evaluate the impact of the improvement plan, also involves a behavioural change intervention developed in stage 4. This will consist of measuring the uptake of occupational health and safety measures in both the intervention and control groups before the intensified behavioural change intervention is introduced, and again after the intervention has been introduced. The selection of the intervention and control groups will be decided after establishing the actual number of de-sludging operators in the study cities.

### Sample size and sampling procedure

The sample size will be calculated using the Kish Leslie formula of cross-sectional studies (Kish, 1965). The researchers assume a prevalence of use of PPE of 88.1% [19], a 5% margin of error, and a 95% confidence interval which yields a sample size of 159 de-sludging operators. Considering a design effect of 2.0 to cater for intra-cluster correlation and an increase in the sampling error [20], and a 10% non-response rate, a total of 356 de-sludging operators will be recruited into the study. The sample size for laboratory samples for intestinal helminth infestation will also be calculated using the Kish Leslie formula of cross-sectional studies (Kish, 1965). This will consider a prevalence of intestinal helminth infestation of 4.5% [19], a margin of error of 5%, and a 95% confidence interval. After adjusting for a 10% non-response rate, there will therefore be a total number of 73 samples required for the assessment of intestinal helminth infestation. Before data collection, the total number of registered de-sludging operators in the 11 cities will be established, after the proportionate sample is obtained. Simple random sampling with replacements will be used in the selection of participants for the quantitative approach. A list of eligible participants will be obtained from the respective city authorities or de-sludging operators’ associations. Computer-generated random numbers (using the EXCEL Random number generator) will be used to randomly select de-sludging operators who will participate in this study. Purposive sampling will also be used in the selection of study participants for the focus group discussions (FGDs) and in-depth interviews (IDIs).

### Data collection methods and tools

A quantitative approach will be used to establish the prevalence of occupational hazards, knowledge, attitude and uptake of OHS measures, and the HRQoL of de-sludging operators. A structured questionnaire will be comprised to elicit information on socio-demographic characteristics, exposure to occupational hazards, self-reported work-related accidents and diseases, as well as uptake of safety measures.

A qualitative approach will be used to explore mechanisms of coping with occupational hazards, and facilitators and barriers to the promotion of OHS of de-sludging operators. A total of 11 FGDs and 15 IDIs will be conducted among purposively selected de-sludging operators to understand the exposure of hazards and coping mechanisms. In addition, 15 key informant interviews will be conducted among purposively selected employers, including city authorities’ representatives, to understand the barriers and facilitators of the provision and implementation of OHS measures among de-sludging operators. Data collection tools have been developed after a thorough review of the literature.

### Stool sample collection and laboratory analysis

In order to characterise biological hazards, stool samples will be analysed. Study participants will be provided with stool containers to sample a day before the survey. Participants will then be requested to provide their first fresh stool passed in the morning of the sample collection day and will also be cautioned not to mix the stool sample with urine or water. Stool samples will be given unique identification numbers and transported for laboratory analysis within 4 hours of collection. While in the laboratory, samples will be subject to the Kato-Katz technique (duplicate thick smears, using standard 41.7 mg templates) and a formalin-ether concentration technique (FECT) as described by [19, 21]. The helminths of interest will include *Ascaris lumbricoides*, Hookworm, and *Trichuris trichiura*.

### Study variables

The main outcome of interest will be the uptake of OHS measures such as the use of PPE and regular health check-ups. The supplementary outcomes will include health-related outcomes such as the prevalence of musculoskeletal disorders, stigma and intestinal helminth infestation, and the HRQoL.

### Measurement of the outcome variables

#### Uptake of OHS measures

During the assessment of uptake of OHS measures, a team of environmental and occupational health experts will undertake a risk assessment to identify and define the hazards that study participants are exposed to during de-sludging operations, as well as the required OHS measures. Afterwards, study participants will be observed to establish the utilization of PPE and their history of medical check-ups. Those practising the required actions (PPE use and medical check-ups) will be considered to consider OHS measures.

#### Prevalence of musculoskeletal disorders

The history of musculoskeletal disorders will be assessed by obtaining information on injury and health complaints among the study participants. The study will adapt the Nordic Musculoskeletal Questionnaire [22, 23]; questions are developed by Okello, Wafula (24) to establish musculoskeletal injuries and health complaints. A “yes” response to complaints from duty will be used to ascertain the prevalence of musculoskeletal disorders.

#### Prevalence of intestinal helminth infestations

Stool samples will be collected by the research team and transported to the laboratory on ice within 4 hours for quantification of helminth among de-sludging operators. Stoll’s technique will be used to detect helminth eggs and helminth in the stool samples. Prevalence of helminth infection will be based on the WHO recommendations i.e. light, moderate, and heavy [25] (Table 1). This approach has previously been used by other scholars to quantify the intensity of helminth infection [19, 21].

**Table 1:**
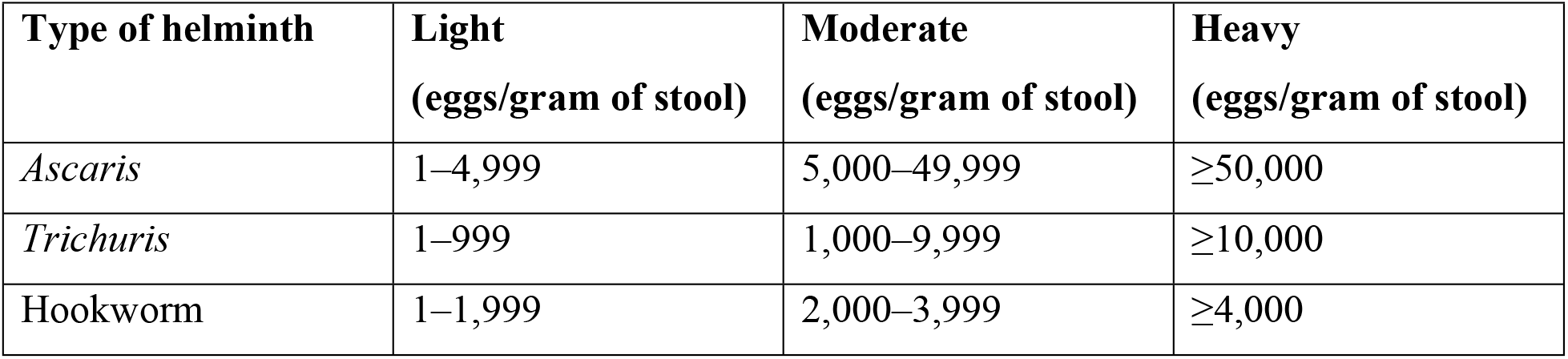
Classification of helminth infection intensities based on Kato-Katz faecal egg counts.

| Type of helminth | Light<br>(eggs/gram of stool) | Moderate<br>(eggs/gram of stool) | Heavy<br>(eggs/gram of stool) |
| --- | --- | --- | --- |
| *Ascaris* | 1–4,999 | 5,000–49,999 | ≥50,000 |
| *Trichuris* | 1–999 | 1,000–9,999 | ≥10,000 |
| Hookworm | 1–1,999 | 2,000–3,999 | ≥4,000 |

#### Measurement of stigma

Stigma will be measured using the social impact scale (SIS), a widely used 24-item measure of stigmatization [26-28]. This will assess 4 domains, including social rejection (9 items), financial insecurity (3 items), internalized shame (5 items), and social isolation (7 items). For each item, participants will indicate whether they agree or disagree on a four-point Likert scale ranging from ‘strongly agree’ to ‘strongly disagree’. Each SIS item/statement will be rated from ‘1’ (strongly disagree) to ‘4’ (strongly agree), with total scores ranging from 24 to 96. Individuals with higher scores will be considered to be experiencing severe stigma.

#### Measurement of HRQoL

The HRQoL will be assessed using a tool adopted from two tools namely: the Center for Disease Control HRQoL tool known as CDC HRQoL -14 [29] and the EQ -5D Europol tool [30, 31]. The adopted tool will entail descriptive questions and a Visual Analog Scale (VAS). The VAS is a self-rated health tool visualized on a 20 cm vertical, visual analogue scale with endpoints labelled ‘the best health one can imagine’ (‘100’) and ‘the worst health one can imagine’ (‘0’). Descriptive questions will be asked to the participants to report their health state in real-time by ticking the most appropriate answer. The results will then be scored and a new variable for HRQoL will be formed. Participants who score greater than or equal to 60% will have a high HRQoL, <60%-50% will have moderate HRQoL while those <50% will have a low HRQoL.

### Measurement of independent variables

#### Prevalence of exposure to occupational hazards

Classification of occupational hazards will be based on the International Occupational Hazards Database of the International Labour Organization Occupational Safety and Health Information Centre [32]. The research will establish the prevalence of exposure to occupational hazards by asking study participants how frequently they were exposed to occupational hazards in the last 30 days. The frequency of exposure will be coded as 0=Never, 1=Rarely, 2=Sometimes, and 3=Always. Besides the self-reports used to establish the prevalence of exposure to occupational hazards, microbial analysis of environmental samples will also be conducted to establish the presence of *Total coliforms* and *E. coli*. Hand rinsates and surface swabs will be obtained through sampling gloves, steering wheels of vacuum trucks, mobile phones, and gumboots. A pre-moistened swab (Enviromax) over an area of 100 cm^2^ will be used. All environmental samples will aseptically be collected and placed in a sterile Whirl Pak bag (Nasco, Modesto, CA), filled with 500 ml of distilled water. Transportation of all environmental samples to the laboratory for analysis will occur within 4 hours, and on ice to reduce the risk of degradation [33, 34]. At the laboratory, swabs will be washed in 10 ml of Phosphate-buffered saline by vortexing, followed by 10-fold serial dilutions. The samples will be tested for *Total coliforms* and *E. coli* using the membrane filtration technique and Chromocult agar. These samples will be incubated for 48 hours at 37 ± 0.5 °C for total coliforms and 24 hours at 44 ± 0.5 °C for *E. coli*.

#### Other independent variables

Age will be collected as a continuous variable and categorized accordingly at analysis. Sex will be collected as a binary variable with responses including male and female. Education level will be categorized as no formal education, for those who have never attended school; primary, if respondents have only attained primary education; secondary for respondents who have only attained or completed secondary education; and tertiary if a respondent attained education above secondary level. Marital status will be collected as a categorical variable including single for respondents not in union; married/cohabiting for the respondents in union/living together; divorced/separated for respondents that have divorced or separated from their spouses; and widowed for those who have lost spouses.

### Quality assurance and control measures

A team of environmental and occupational health scientists with a minimum of a bachelor’s degree will be recruited as research assistants. These research assistants will undergo a 7-days training to get acquainted with the study protocol, and the ethical issues pertaining to the study. All data collection tools will be pretested and back-translated into the local dialect. Pretesting of the data collection tools will be done among de-sludging operators in Agago town council since these share similar characteristics as those in our study setting. Quantitative data will be entered using the KoboCollect mobile application which will include in-built validation mechanisms and skip patterns. Research assistants will be supervised during the entire data collection period. Daily debrief meetings will be held to identify challenges that may arise during the data collection process and address them accordingly.

### Data analysis and management

#### Quantitative data

Quantitative data will be collected using the KoboCollect mobile application, which will be pre-installed on mobile devices. This will allow for real-time data collection, entry, and preliminary analyses [35]. Afterwards, data will be downloaded in Microsoft Excel to identify and review outliers, code open-ended data as well as cleaned to check for consistency and structural errors. The data cleaning process will also enable the data management team to perform quality assurance checks. Quantitative data will be analysed using STATA version 15. Descriptive analysis will be conducted to summarize categorical and continuous data while logistic regression or modified Poisson regression will be used for inferential statistics. Logistic regression will be used to measure the strength of association between the independent variables and rare outcomes (prevalence less than 10%) [36, 37]. For outcomes whose prevalence will exceed 10%, modified Poisson regression analysis with robust standard errors will be used to report prevalence ratios [38, 39].

#### Qualitative data

Qualitative data will be collected by two research assistants (i.e. an interviewer and a note-taker) at all times. All interviews will be digitally recorded to enable the research assistants to focus on listening, probing, following, and maintaining eye contact with the interviewees. The audio recordings will be transcribed verbatim by two experienced transcribers. Qualitative data will be managed in NVIVO 12.0. Data coding will be done by two qualitative experts (T.S and A.N). A blended deductive and inductive approach will be used during the qualitative analysis. Illustrative quotes will be used during reporting of qualitative findings.

### Dissemination of study findings

Dissemination workshops will be organized (either physically or virtually) with various stakeholders such as respective city authorities, Water Aid Uganda, Uganda Gulpers Association, Private Emptiers Association Uganda, and Kampala Emptiers Association. Copies of the written report will be shared with the relevant stakeholders. Policy briefs will also be drafted based on the findings from the study; as well as electronic/virtual dissemination platforms such as social media (Twitter/Facebook/ LinkedIn) and blogs. Manuscripts will also be written and submitted to peer-reviewed journals for publication.

### Community engagement plan

This study was conceptualized after consultation with the study community (in this case, de-sludging operators). In order to increase ownership of the study interventions and findings, a representative of the de-sludging operators has been named as one of the research team members. During the implementation of the study, de-sludging operators with a minimum diploma qualification will be involved in the data collection process, thus increasing ownership of the study findings. The researchers plan to engage the representatives of the de-sludging operators in the validation of the study findings and review of policy briefs to ensure the problems that affect them are brought forward to the policymakers. The researchers also plan to engage with respective city authorities and relevant stakeholders in the development of the study protocol, design of data collection tools, and validation of study findings.

### COVID-19 risk mitigation plan

Field data collection during this period of COVID-19 poses a risk of infection with the coronavirus to the study participants and research assistants. In order to minimize this risk, a risk mitigation plan (RMP) has been developed which will be followed strictly by the study participants and research assistants. The RMP highlights standard precautions to be followed during (a) transportation of field staff; (b) training of research assistants; (c) field data collection, and (d) after field data collection is completed. In general, all participants will be strongly encouraged to wear a face mask during all interactions with the research team and fellow participants; and all field teams will carry a hand sanitiser that will be used for hand hygiene before handling any paperwork. Research assistants will be required to hand-sanitize before commencing interviews; keep adequate social distance during interviews and in all situations where this is warranted, and wear a face mask while in the field at all times.

## Discussion

The main objective of this study is to use the SSP tool to improve the OHS of de-sludging operators in Uganda. Quantitatively, the study aims to (1) establish the prevalence of occupational hazards and associated health outcomes among de-sludging operators, and (2) establish the knowledge, attitude and uptake of OHS measures. Qualitatively, the study aims to understand the facilitators and barriers to the promotion of occupational health and safety of desludging operators.

De-sludging operators in developing countries are exposed to a multitude of physical, chemical, biological, and psychosocial occupational hazards, which eventually affect their HRQoL. Although not quantified, sanitation workers, including de-sludging operators in Uganda, work under poor conditions which expose them to (1) physical hazards such as falls, slips, cuts, and impact by flying objects, (2) chemical hazards such as detergents and toxic gases, (3) biological hazards such as bacteria and viruses contained in excreta and wastewater, (4) ergonomic hazards such as repetitive motions/posture and frequent lifting, and (5) psychosocial hazards such as verbal abuse, and stigma and discrimination [6]. Exposure to these hazards is often exacerbated by the dilapidated and confined nature of the sanitation facilities, inaccessible neighbourhoods, and negative community perceptions of de-sludging operators.

Research activities will start with the identification and assembly of key stakeholders involved in OHS of de-sludging operators. These activities will include among others the local authorities’ staff, de-sludging operators and their association leaders, policymakers, and the communities served. Together with the key stakeholders, the study will use qualitative methods to understand the existing OHS conditions and mitigation measures. At this stage, priority areas for intervention will be established, and set objectives, targets, and strategies for monitoring progress. Stakeholder engagement is critical for increasing ownership and sustainability of the proposed interventions [40, 41]. Setting objectives, targets and a monitoring system is important in tracking changes in uptake of OHS measures and will provide the basis for evaluation of the interventions [42-44].

This research will use a team of environmental health experts to undertake an assessment of the sanitation chain starting with the containment sites, followed by conveyance/transportation and disposal. Undertaking this assessment will enable the research team to understand the array of hazards that de-sludging operators are exposed to and the existing mitigation measures. At this stage, the team of environmental health experts will document hazards, hazardous events, exposure routes, and the likelihood of occurrence of the hazardous events. A risk analysis matrix will be used to rank the risks associated with each hazardous event and develop appropriate mitigation measures [45].

During the data collection phase, the prevalence of exposure to physical, ergonomic, biological, and psychosocial hazards, and utilisation of PPE along the sanitation chain will be established. The potential hazards among desludging operators include dust, vapour, gas, oxygen deficient atmospheres, noise from various sources, electrostatic build-up, slipping, cuts, and punctures, falling objects, metal, and chemical splash, abrasion, and repetitive motions [5]. The researchers will use a modified version of the standardized Nordic and Washington state risk factor checklist and the upper limb Core QX checklist [23, 46, 47] to establish the prevalence of musculoskeletal disorders. These tools have previously been used to study musculoskeletal disorders among other sanitation workers [48].

Self-reports will be used to establish the prevalence of exposure to physical, chemical, and biological hazards, and uptake of occupational health and safety measures. Although self-reports can be characterised by biases, these have been used to get insights into exposure to occupational hazards and uptake of prevention measures among individuals involved in the delivery of sanitation services [49]. Uptake of occupational health and safety measures, such as PPE along the sanitation chain, has been reported to mitigate occupational health risks among sanitation workers, including those engaged in desludging [5, 49]. The common PPE used among sanitation workers include gloves, respiratory masks, gas monitors and gumboots, goggles, helmets, jackets, and safety cones [5].

Understanding facilitators and barriers to the promotion of OHS is necessary to inform the design of appropriate interventions geared toward effective and sustainable OHS [50]. A review of literature reveals that barriers and facilitators to OHS occur across all levels (extra organizational, organizational, worker, and program levels) due to limited resource availability [50]. Factors such as training de-sludging operators on OHS, supervision, policy and regulations, and enforcement, influence uptake and promotion of OHS measures [49].

Following the establishment of mitigation measures and understanding of the facilitators and barriers to the promotion of OHS, the research team will guide the key stakeholders in developing an improvement plan which will consider the status quo, available resources, and the policy environment. This stage will provide an opportunity for stakeholders to identify their strengths and weaknesses in relation to OHS. It will also act as a roadmap to achieving OHS targets and goals, as documented by earlier scholars [51, 52].

### Strengths and limitations

This may be the first study to use the SSP tool to improve the occupational health and safety (OHS) of de-sludging operators in Uganda. However, since the researchers plan to use self-reports to assess some variables, the study is subject to biases such as the social desirability bias and recall. The researchers plan to recruit experienced research assistants and thoroughly train them to minimize these biases. In addition, interviewees will be allowed sufficient time during interviews for adequate recall.

## Conclusion

This study will generate evidence of the prevalence of occupational hazards and associated health outcomes among de-sludging operators. This extends to increasing the knowledge, attitude and uptake of OHS measures and the facilitators and barriers in the promotion of the occupational health and safety of desludging operators. This evidence will be used to inform policy and practice.

## Data Availability

o datasets were generated or analysed during the current study. All relevant data from this study will be made available upon study completion.

## Abbreviations

GDP: Gross Domestic Product
GKMA: Greater Kampala Metropolitan Area
HBM: Health Belief Model
HRQoL: Health-Related Quality of Life
OHS: Occupational Health and Safety
PPE: Personal Protective Equipment
SSP: Sanitation Safety Planning

## Declarations

### Ethics approval and consent to participate

Ethical approval was obtained from the Makerere University School of Public Health Research Ethics Committee. The study was registered with the Uganda National Council of Science and Technology. Administrative clearance will be obtained from the respective City authorities and private companies. Written informed consent will also be obtained from the study participants and all the information provided by the study participants will be kept confidential.

### Consent to publish

Not applicable

### Availability of data and materials

The datasets analysed during the current study are available from the corresponding author upon reasonable request.

### Authors’ contributions

TS, AN and JBI obtained funding for the proposed study. TS, AN, SF, RKM, WKK, AT, DN, FN, CK, JSM, BNT, AAH, and JBI participated in the conceptualization and development of this protocol. All authors read and approved this manuscript before submission to this journal.

## Acknowledgement

We appreciate the leaders of the de-sludging operators association and the city authorities that informed the design of the proposed study.

